# Clinical Outcomes of Pulmonary Vein Isolation Versus Antiarrhythmic Drugs as First-Line Therapy for Atrial Fibrillation: A Propensity Score-Matched Analysis

**DOI:** 10.1101/2025.05.23.25328263

**Authors:** Yong Hao Yeo, Aravinthan Vignarajah, Hermon Kha Kin Wong, Nishanthi Vigneswaramoorthy, Jian Liang Tan, Beeletsega T. Yeneneh, Komandoor Srivathsan, Justin Lee, Dan Sorajja

## Abstract

**Background:** Pulmonary vein isolation (PVI) has increasingly demonstrated superiority over antiarrhythmic drugs (AAD) for rhythm control in atrial fibrillation (AF). However, large-scale, long-term, real-world studies comparing these two therapies as first-line AF management remain limited.

**Methods:** Using the TriNetX network, we identified patients (≥18 years old) with AF between 2012 and 2019. Patients were categorized into two cohorts: PVI vs. AAD as first-line therapy for AF. Patients were followed for 5 years, with the primary outcome being a composite of all-cause death, all-cause hospitalization, and heart failure exacerbation. Secondary outcomes included ischemic stroke and major bleeding events (intracranial bleeding/ gastrointestinal bleeding). Subanalyses were performed in the paroxysmal and persistent AF cohorts, respectively.

**Results:** Among 342,230 eligible patients, 2,638 patients (mean age 64.3 ± 10.6 years) who underwent PVI and 2,638 patients (mean age 64.2 ± 13.1 years) who had AAD as first-line therapy for AF had similar propensity scores and were included in the analysis. At 5-year follow-up, the PVI group had a lower risk of the primary composite outcome compared to the AAD group (42.0% vs. 51.1%; HR 0.76; 95% CI, 0.71–0.83; P < 0.01). They also had lower risk of all-cause mortality (4.1% vs. 7.7%; HR 0.53; 95% CI, 0.42–0.67; P < 0.01), all-cause hospitalization (35.1% vs. 42.2%; HR 0.77; 95% CI, 0.71–0.84; P < 0.01), and heart failure exacerbation (21.0% vs. 24.3%; HR 0.85; 95% CI, 0.76–0.95; P < 0.01. Ischemic stroke (6.1% vs. 6.7%; HR 0.90; 95% CI, 0.73–1.12; P = 0.34), and major bleeding event (4.3% vs. 5.3%; HR 0.80; 95% CI, 0.62–1.02; P = 0.08) were similar between groups. Similar outcomes were seen in both the paroxysmal and persistent AF cohorts.

**Conclusion:** After a 5-year follow-up period, PVI was associated with better clinical outcomes than AAD as first-line therapy for AF.

## INTRODUCTION

Atrial fibrillation (AF) is the most prevalent cardiac arrhythmia, significantly contributing to global morbidity and mortality.^1–3^ Effective rhythm control is essential to mitigate the risks associated with AF, including stroke, heart failure, and overall cardiovascular burden. Current rhythm control strategies primarily involve antiarrhythmic drugs (AAD) and catheter-based pulmonary vein isolation (PVI). Over the past two decades, PVI has emerged as a cornerstone of non-pharmacological AF management, demonstrating superior efficacy in maintaining sinus rhythm compared to AAD in selected populations.^4^

Several landmark trials, including MANTRA-PAF, RAAFT-2, STOP AF, and EARLY-AF, have established the benefits of PVI over AAD in patients with paroxysmal AF, leading to guideline recommendations advocating for PVI as a first-line therapy in this subset.^5–9^ However, despite these advances, data on the long-term effectiveness of PVI as first-line therapy in real-world settings remain limited. Furthermore, while PVI has been extensively studied in paroxysmal AF, its role in persistent AF is less well defined, necessitating further investigation into its comparative efficacy in this subgroup.

To address these gaps, we conducted a large-scale retrospective analysis comparing the long-term outcomes of PVI versus AAD as first-line therapy in patients with AF. Additionally, we performed a subanalysis to assess the differential impact of these strategies in patients with paroxysmal versus persistent AF. Our study aims to provide a more comprehensive understanding of the effectiveness of these rhythm control approaches, potentially guiding clinical decision-making and informing future guideline recommendations.

## METHODS

### Data Source

We used TriNetX, a global federated health research network providing access to statistics on electronic medical records (diagnoses, procedures, medications, laboratory values, genomic information) from approximately 130 million patients in 100 large Healthcare Organizations, predominantly in the United States. As a federated network, TriNetX received a waiver from Western IRB since only aggregated counts, statistical summaries of de-identified information, but no protected health information is received, and no study-specific activities are performed in retrospective analyses.

### Study Population and Design

We used ICD-10-CM codes to identify patients aged 18 and older with AF (ICD-10-CM code I48) from January 1, 2012 to January 1, 2019. The cohort was divided into two groups: those receiving PVI and those receiving AAD as first-line therapy for rhythm control of AF. AAD included amiodarone, dronedarone, dofetilide, sotalol, and flecainide. Subanalyses were performed in the paroxysmal and persistent AF cohort, respectively.

### Study Outcomes

The primary outcome was a composite of all-cause mortality, all-cause hospitalization and heart failure exacerbation. Secondary outcomes included ischemic stroke and major bleeding event (intracranial or gastrointestinal bleeding). We analyzed clinical outcomes that occurred between 90 days and 5 years following rhythm control therapy.

### Statistical Analysis

Continuous variables were presented as mean ± standard deviation (SD) and compared using independent-sample t-tests, while categorical variables were expressed as counts and percentages (n, %) and analyzed with chi-square tests. To account for baseline differences between cohorts, a 1:1 propensity-score matching (PSM) approach was applied using a greedy nearest-neighbor method with a caliper of 0.1 pooled standard deviations. All variables listed in Table 1 (including patient demographics, medical comorbidities, medications, and lab values) were incorporated into the PSM model. Variables with a P-value greater than 0.05 were considered well-matched. Hazard ratios were calculated to assess the relationship between GLP-1 RA use versus control and the outcomes, with 95% confidence intervals (CI) used to evaluate the precision and statistical significance of these ratios.

**Table 1.** Baseline Characteristics of Patients with Pulmonary Vein Isolation vs. Antiarrhythmic Drugs as First-Line Therapy for Atrial Fibrillation (Before and After Propensity Score Matching).

|  | Before Propensity Score Matching |  |  | After Propensity Score Matching |  |  |
| --- | --- | --- | --- | --- | --- | --- |
|  | PVI, n (%) | AAD, n (%) | P-Value | PVI, n (%) | AAD, n (%) | P-Value |
| No. of patients | 2,639 | 339,591 |  | 2,638 | 2,638 |  |
| **Patient Demographics** |  |  |  |  |  |  |
| Age, mean (SD), y | 64.3 +/- 10.6 | 69.7 +/- 12.1 | <0.01 | 64.3 +/- 10.6 | 64.2 +/- 13.1 | 0.75 |
| Female | 787 (29.8) | 136,152 (40.1) | <0.01 | 787 (29.8) | 783 (29.7) | 0.90 |
| Race or Ethnic Groups |  |  |  |  |  |  |
| *White* | *2,190 (83.0)* | *260,629 (76.7)* | *<0.01* | *2,189 (83.0)* | *2,210 (83.8)* | *0.44* |
| *Hispanic or Latino* | *73 (2.8)* | *11,512 (3.4)* | *0.08* | *73 (2.8)* | *77 (2.9)* | *0.74* |
| *Black or African American* | *81 (3.1)* | *27,287 (8.0)* | *<0.01* | *81 (3.1)* | *59 (2.2)* | *0.06* |
| *Asian* | *38 (1.4)* | *12,742 (3.8)* | *<0.01* | *38 (1.4)* | *35 (1.3)* | *0.72* |
| **Concomitant Medical Comorbidities** |  |  |  |  |  |  |
| Chronic Kidney Diseases |  |  |  |  |  |  |
| *Chronic Kidney Disease Stage 3* | *122 (4.6)* | *37,153 (10.9)* | *<0.01* | *122 (4.6)* | *115 (4.4)* | *0.64* |
| *Chronic Kidney Disease Stage 4* | *12 (0.5)* | *12,138 (3.6)* | *<0.01* | *12 (0.5)* | *19 (0.7)* | *0.21* |
| *End-Stage Kidney Disease* | *19 (0.7)* | *14,804 (4.4)* | *<0.01* | *19 (0.7)* | *20 (0.8)* | *0.87* |
| Chronic Obstructive Pulmonary Disease | 148 (5.6) | 52,920 (15.6) | <0.01 | 148 (5.6) | 149 (5.6) | 0.95 |
| Coronary Artery Disease | 813 (30.8) | 157,429 (46.4) | <0.01 | 813 (30.8) | 770 (29.2) | 0.20 |
| Diabetes Mellitus | 450 (17.1) | 101,065 (29.8) | <0.01 | 450 (17.1) | 432 (16.4) | 0.51 |
| Hyperlipidemia | 1,127 (42.7) | 177,559 (52.3) | <0.01 | 1,126 (42.7) | 1,064 (40.3) | 0.08 |
| Hypertension | 1,522 (57.7) | 216,871 (63.9) | <0.01 | 1,521 (57.7) | 1,471 (55.8) | 0.17 |
| Malignancy | 437 (16.6) | 84,383 (24.8) | <0.01 | 437 (16.6) | 391 (14.8) | 0.08 |
| Obesity | 577 (21.9) | 67,173 (19.8) | <0.01 | 576 (21.8) | 568 (21.5) | 0.79 |
| Thyroid Disorders | 371 (14.1) | 66,866 (19.7) | <0.01 | 371 (14.1) | 332 (12.6) | 0.11 |
| **Medications** |  |  |  |  |  |  |
| Anticoagulants |  |  |  |  |  |  |
| *Apixaban* | 807 (30.6) | 54,561 (16.1) | <0.01 | 807 (30.6) | 839 (31.8) | 0.34 |
| *Dabigatran* | 279 (10.6) | 13,696 (4.0) | <0.01 | 278 (10.5) | 246 (9.3) | 0.14 |
| *Rivaroxaban* | 631 (23.9) | 40,111 (11.8) | <0.01 | 630 (23.9) | 592 (22.4) | 0.22 |
| *Warfarin* | 604 (22.9) | 88,395 (26.0) | <0.01 | 603(22.9) | 567 (21.5) | 0.23 |
| Anti-Diabetic Medications |  |  |  |  |  |  |
| *Glipizide* | 44 (1.7) | 13,407 (3.9) | <0.01 | 44 (1.7) | 40 (1.5) | 0.66 |
| *Insulin* | 227 (8.6) | 113,780 (33.5) | <0.01 | 227 (8.6) | 194 (7.4) | 0.09 |
| *Metformin* | 202 (7.7) | 37,297 (11.0) | <0.01 | 202 (7.7) | 199 (7.5) | 0.87 |
| *Sitagliptin* | 33 (1.3) | 9,397 (2.8) | <0.01 | 33 (1.3) | 36 (1.4) | 0.72 |
| Beta-blocker | 1,644 (62.3) | 251,844 (74.2) | <0.01 | 1,643 (62.3) | 1,586 (60.1) | 0.11 |
| Diuretics | 1,309 (49.6) | 196,336 (57.8) | <0.01 | 1,309 (49.6) | 1,240 (47.0) | 0.06 |
| Digoxin | 215 (8.1) | 49,965 (14.7) | <0.01 | 215(8.2) | 181(6.9) | 0.08 |

Survival analysis was performed by plotting Kaplan-Meier curves and comparing the two cohorts using log-rank tests. Statistical significance was set at a two-sided P value of <0.05. All analyses were conducted using the TriNetX online platform and R for statistical computing.

## RESULTS

### Study Population

Our study included 342,230 patients aged ≥18 years with AF from January 1, 2012, to January 1, 2019. 2,639 patients (mean age 64.3 ± 10.6 years, 29.8% female) received PVI as first-line therapy, while 339,591 patients (mean age 69.7 ± 12.1 years, 40.1% female) were treated with AAD as first-line therapy. **Figure 1** demonstrates the patient selection process. Among the patients receiving AAD as first-line therapy, amiodarone is the most commonly prescribed AAD (231,492, 66.8%), followed by sotalol (45,718, 13.2%), flecainide (41,992, 12.1%), dronedarone (21,699, 6.3%), and dofetilide (18,138, 5.2%).

**Figure 1.**
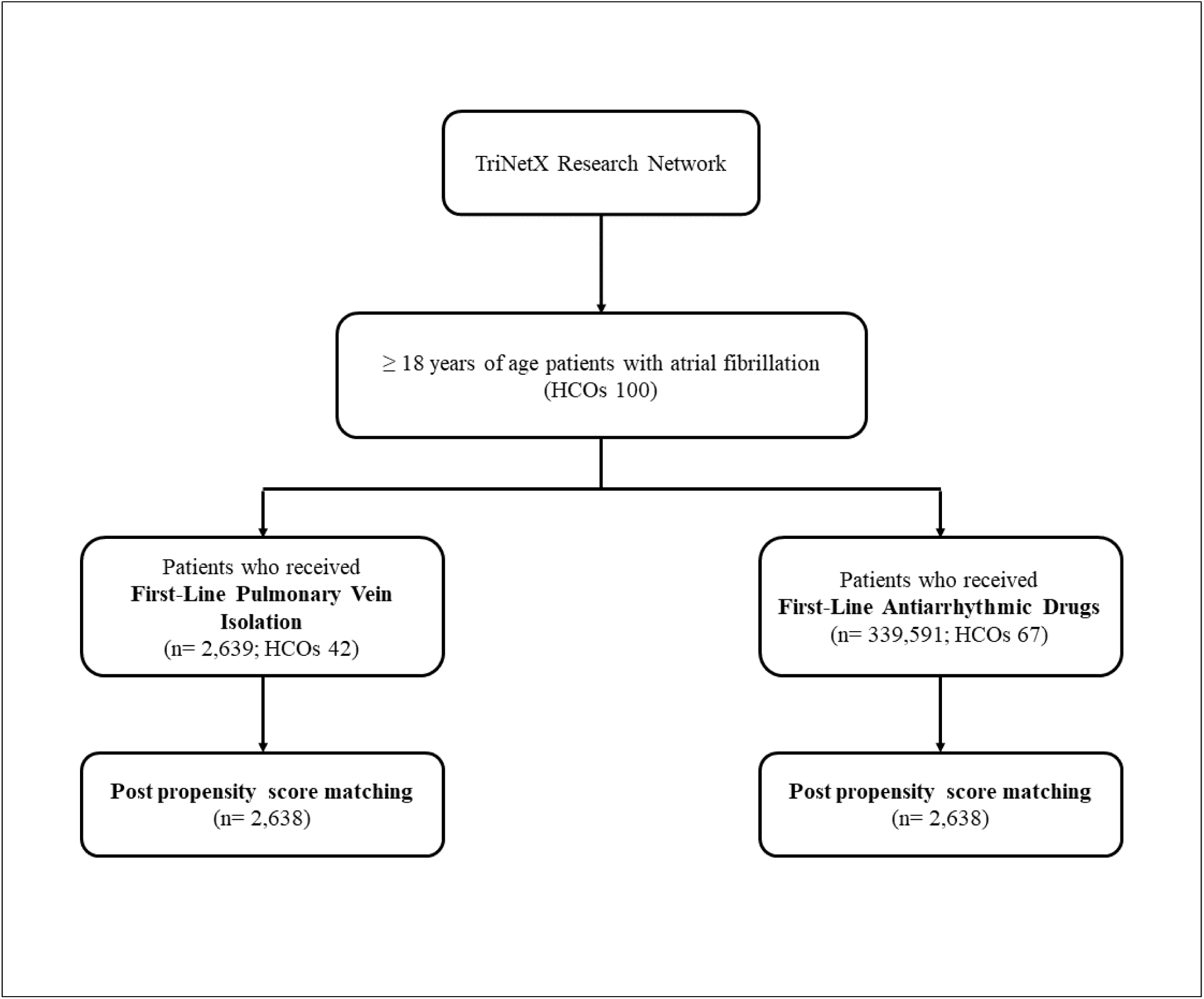
Patient Selection Process (CONSORT Diagram).

Patients’ baseline characteristics are summarized in **Table 1**. Before PSM, patients receiving PVI as first-line therapy were younger and had a higher proportion of males compared to those receiving AAD. The majority of patients in both groups were White. Patients in the PVI group had a lower prevalence of most concomitant medical conditions, except for obesity. After PSM, 2,638 matched patients were included in each group for further analysis.

### Study Outcomes

**Table 2** presents the percentages of outcomes in both groups of patients. At 5-year follow-up, the PVI group had a lower risk of the primary composite outcome compared to the AAD group (42.0% vs. 51.1%; HR 0.76; 95% CI, 0.71–0.83; P < 0.01). Patients undergoing PVI experienced a lower risk of all-cause mortality (4.1% vs. 7.7%; HR 0.53; 95% CI, 0.42–0.67; P < 0.01), all-cause hospitalization (35.1% vs. 42.2%; HR 0.77; 95% CI, 0.71–0.84; P < 0.01 and heart failure exacerbation (21.0% vs. 24.3%; HR 0.85; 95% CI, 0.76–0.95; P = <0.01) than those receiving AADs. The risks of ischemic stroke (6.1% vs. 6.7%; HR 0.90; 95% CI, 0.73–1.12; P = 0.34), and major bleeding event (4.3% vs. 5.3%; HR 0.80; 95% CI, 0.62–1.02; P = 0.08) were similar between groups (**Figure 2**). Kaplan-Meier survival curves for all-cause mortality are shown in **Figure 3**.

**Figure 2.**
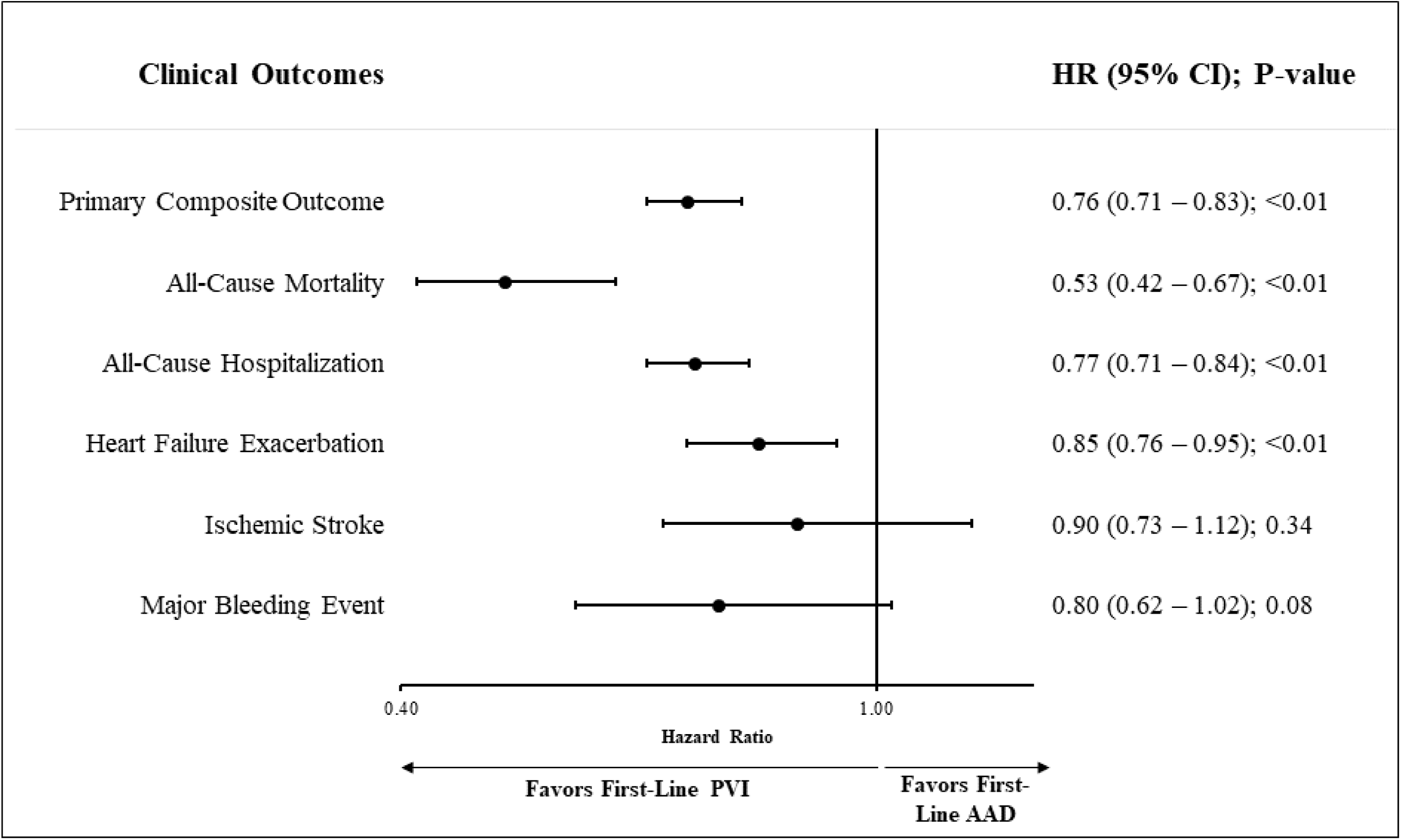
Measures of Association for Clinical Outcomes of Patients with Pulmonary Vein Isolation vs. Antiarrhythmic Drugs as First-Line Therapy for Atrial Fibrillation (Antiarrhythmic Drugs as Reference).

**Figure 3.**
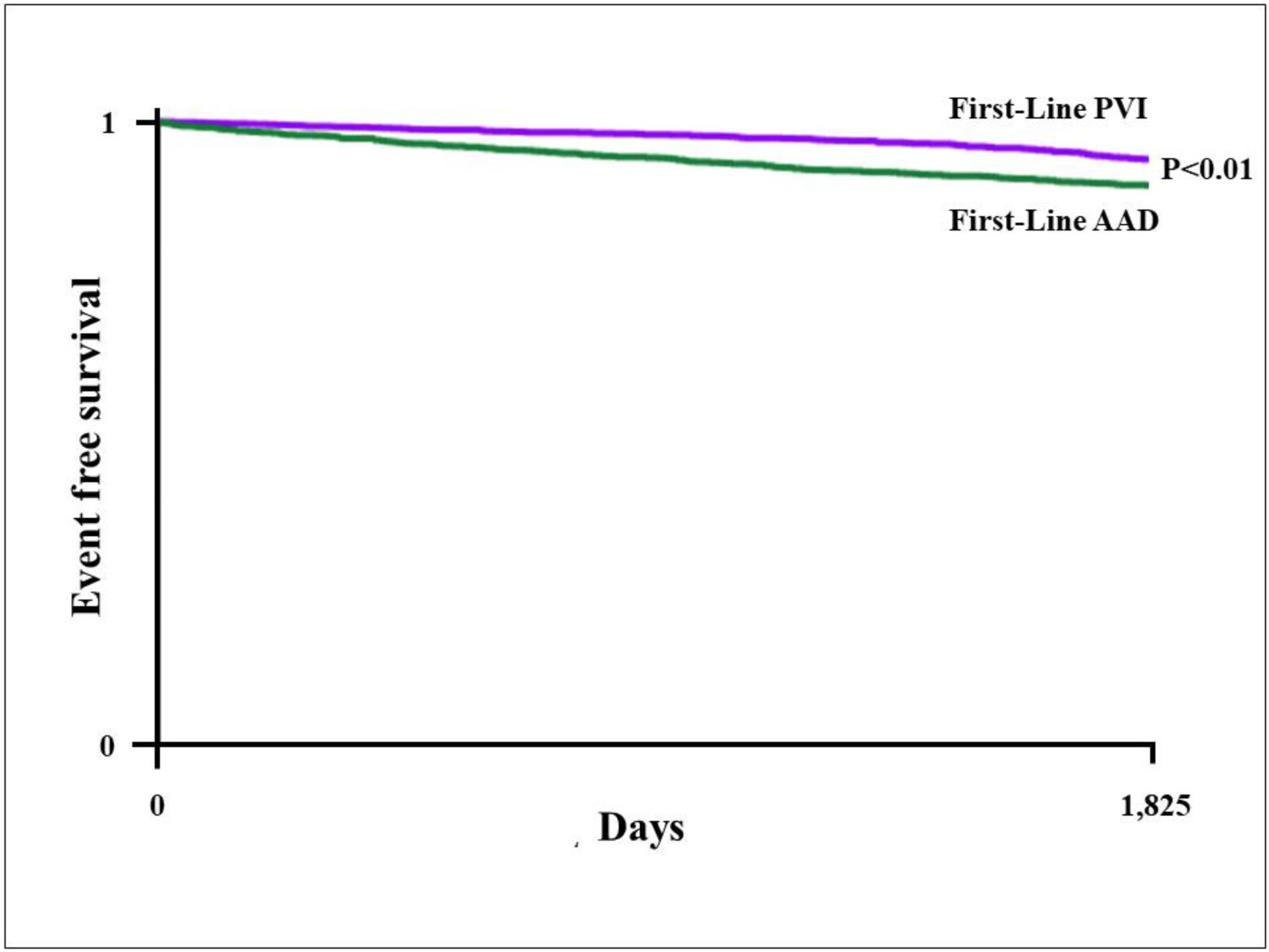
Kaplan-Meier Survival Analyses of All-Cause Mortality for Pulmonary Vein Isolation vs. Antiarrhythmic Drugs as First-Line Therapy for Atrial Fibrillation.

**Table 2.** Clinical Outcomes of Patients with Pulmonary Vein Isolation vs. Antiarrhythmic Drugs as First-Line Therapy for Atrial Fibrillation (Before and After Propensity Score Matching).

|  | **PVI, n (%)** | **AAD, n (%)** | **Hazard Ratio**<br>**(95% CI)**<br>**(AAD as**<br>**reference)** | **P-value** |
| --- | --- | --- | --- | --- |
| **Primary Outcomes** |  |  |  |  |
| Composite Outcome | 1,107 (42.0) | 1,348 (51.1) | 0.76 (0.71 – 0.83) | <0.01 |
| All-Cause Mortality | 108 (4.1) | 202 (7.7) | 0.53 (0.42 – 0.67) | <0.01 |
| All-Cause Hospitalization | 927 (35.1) | 1,112 (42.2) | 0.77 (0.71 – 0.84) | <0.01 |
| Heart Failure Exacerbation | 553 (21.0) | 642 (24.3) | 0.85 (0.76 – 0.95) | <0.01 |

| Secondary Outcomes |  |  |  |  |
| --- | --- | --- | --- | --- |
| Ischemic Stroke | 161 (6.1) | 178 (6.7) | 0.90 (0.73 – 1.12) | 0.34 |
| Major Bleeding Event (Intracranial or Gastrointestinal Bleeding) | 113 (4.3) | 140 (5.3) | 0.80 (0.62 – 1.02) | 0.08 |

### Subcohort Analyses

We conducted a subgroup analysis to compare clinical outcomes between PVI and AAD therapy as first-line treatment in patients with paroxysmal and persistent AF. In the paroxysmal AF cohort, 602 matched patients were included in each group. At 5-year follow-up, PVI was associated with a significantly lower risk of the primary composite outcome compared to AAD therapy (36.2% vs. 46.3%; HR 0.71; 95% CI, 0.60–0.85; P < 0.01). PVI also conferred a lower risk of all-cause mortality (2.5% vs. 5.1%; HR 0.49; 95% CI, 0.27–0.91; P = 0.02), all-cause hospitalization (31.9% vs. 40.9%; HR 0.71; 95% CI, 0.59–0.86; P < 0.01), and major bleeding events (2.7% vs. 5.1%; HR 0.52; 95% CI, 0.28–0.95; P = 0.03). No significant differences were observed between groups in the rates of heart failure exacerbation (11.5% vs. 14.5%; HR 0.79; 95% CI, 0.58–1.09; P = 0.15) or ischemic stroke (5.3% vs. 5.0%; HR 1.08; 95% CI, 0.66–1.78; P = 0.76) (**Supplemental Table 1**).

In the persistent AF cohort, 2,490 matched patients were included in each group. PVI remained significantly associated with a reduced risk of the primary composite outcome (42.9% vs. 51.8%; HR 0.75; 95% CI, 0.69–0.81; P < 0.01), as well as lower risks of all-cause mortality (4.3% vs. 9.8%; HR 0.42; 95% CI, 0.34–0.53; P < 0.01) and all-cause hospitalization (35.7% vs. 42.7%; HR 0.76; 95% CI, 0.69–0.83; P < 0.01). Additionally, unlike the paroxysmal cohort, PVI was associated with a significant reduction in heart failure exacerbations (21.8% vs. 25.5%; HR 0.83; 95% CI, 0.74–0.93; P < 0.01). The incidence of ischemic stroke remained comparable between groups (6.3% vs. 7.0%; HR 0.89; 95% CI, 0.72–1.10; P = 0.29), and the difference in major bleeding events was not statistically significant (4.5% vs. 5.5%; HR 0.81; 95% CI, 0.63–1.04; P = 0.09) (**Supplemental Table 2**).

Kaplan-Meier survival analyses of all-cause mortality in patients receiving PVI vs. AAD as first-line therapy for paroxysmal and persistent AF are illustrated in **Supplemental Figures 1 and 2**, respectively.

## DISCUSSION

This retrospective longitudinal observational study compared first-line rhythm control strategies for AF, demonstrating superior outcomes for PVI compared to AAD. At five-year follow-up, PVI was associated with a lower hazard ratio of the primary composite outcome compared to AAD. PVI was also linked to reduced hazard ratios of all-cause mortality, all-cause hospitalization, and heart failure exacerbation. The risks of ischemic stroke and major bleeding events were similar between the two groups. Similar outcomes were seen in paroxysmal and persistent AF (**Central Illustration**).

**Central Illustration.**
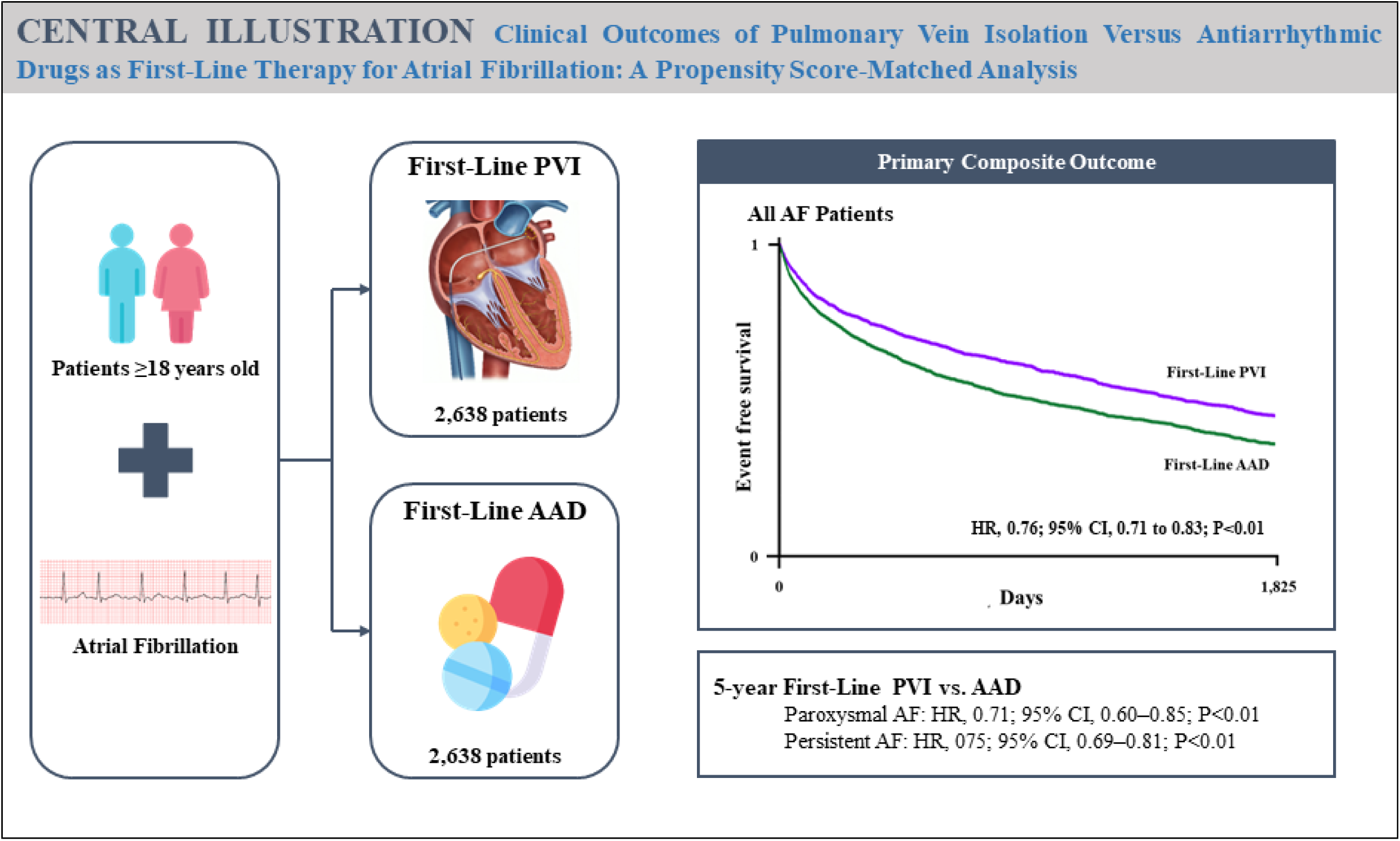
Clinical Outcomes of Pulmonary Vein Isolation Versus Antiarrhythmic Drugs as First-Line Therapy for Atrial Fibrillation.

Our study demonstrated a 47% relative risk reduction in five-year all-cause mortality for the PVI group compared to the AAD group when used as first-line rhythm control therapy for AF. To date, no randomized trials have directly compared PVI and AAD as first-line rhythm control strategies with all-cause mortality as the sole primary endpoint rather than a composite endpoint. Most recent trials reported none or very few deaths, except the MANTRA-PAF trial, which documented seven deaths (2-3%) during its study period.^6,7,10^ Our ability to uniquely compare all-cause mortality rates between these two treatment approaches is attributable to several key factors.

First, given that AF is not typically associated with immediate fatal complications, our extended follow-up period enabled the capture of mortality events that occurred over time. In contrast, most prior trials had a follow-up duration of one year, with RAAFT-2 and EARLY-AF being the exceptions, following patients for up to two and three years, respectively.^8,10,11^ Second, we included a significantly larger and more diverse patient cohort compared to previous trials (5,276 patients in our study versus fewer than 500 in most trials).^8,10,11^ Our study benefits from a large sample size with the statistical power to establish a definitive mortality benefit of PVI as a first-line treatment strategy for AF.

The superiority of PVI in maintaining sinus rhythm likely explains its benefit in reducing heart failure exacerbations, as demonstrated in our study. This finding aligns with a recent meta-analysis by Oraii et al., which focused on AF ablation in patients with heart failure, showing a reduced risk of heart failure events in those with heart failure with reduced ejection fraction.^12^ Similarly, our study demonstrated a substantial reduction in the odds of all-cause hospitalization, suggesting that fewer heart failure events may have indirectly contributed to reduced hospital visits. This reduction in hospitalization odds is consistent with findings from Turagam et al.’s meta-analysis, which included clinical trials assessing catheter ablation versus AAD as first-line therapy for AF.^13^ By decreasing heart failure exacerbations and hospitalizations, PVI not only reduces the healthcare burden but also likely improves patients’ quality of life, a benefit well-documented in recent clinical trials.^6,11^

On the other hand, our study demonstrated comparable risks of ischemic stroke and major bleeding events between the PVI and AAD groups. Previous trials evaluating first-line PVI versus AAD often reported few or no ischemic strokes and major bleeding events, limiting the ability to assess statistical significance due to small sample sizes or low event rates.^5,8,10,11^ In contrast, our results, drawn from a broader and more contemporary patient population, suggest that the stroke-preventive and bleeding risk profiles of the two strategies may be more alike than previously assumed.^14,15^ Theoretically, PVI should reduce stroke risk by improving rhythm control.

However, procedural factors like vascular instrumentation and myocardial injury may alter thromboembolic risk. Therefore, current guidelines recommend continuing anticoagulation for at least three months post-procedure, with longer durations based on individual risk.^16,17^ Despite these recommendations, real-world studies report poor adherence to anticoagulation therapy, even among high-risk patients.^18^ We hypothesize that this complex interplay between rhythm control and anticoagulation patterns may account for our study’s comparable stroke and bleeding rates. However, as our dataset lacks information on post-procedural anticoagulant use, this hypothesis remains unconfirmed and highlights the need for future studies examining anticoagulation patterns following rhythm control.

The 2023 AHA/ACC/HRS AF guideline update reaffirmed catheter ablation as a Class I recommendation for first-line rhythm control therapy in young patients with paroxysmal AF, consistent with the 2017 expert consensus statement.^16,17^ However, the 2023 guideline does not extend this Class I recommendation to persistent AF.^17^ In contrast, the 2017 expert consensus assigned a Class IIa or IIb recommendation for catheter ablation as a first-line therapy in persistent or long-standing AF.^16^ Our findings demonstrated that the benefits of PVI as first-line rhythm control therapy, including lower risks of all-cause mortality, all-cause hospitalization, and heart failure exacerbation, may also apply to patients with persistent AF. These results provide valuable insights into the outcomes of PVI and AAD across AF types.

## STRENGTHS AND LIMITATIONS

Our study has several key strengths. First, it utilizes a large, real-world dataset, providing a comprehensive evaluation of clinical outcomes over a substantial 5-year follow-up period. Second, using propensity score matching allows for a robust comparison of clinical outcomes between patients receiving PVI and those receiving AAD, with minimal confounding factors. Nevertheless, this study has several limitations to consider when interpreting the results. First, the results rely heavily on the clinicians’ accurate documentation. Secondly, the database lacks a few granular information, including the exact burden of AF, procedural details, and causes of death. Therefore, we could not adjudicate the cause of death to identify the exact reason for the mortality benefit of PVI. Thirdly, there is limited diversity in the demographic groups. Certain demographic groups, such as underrepresented minorities, may not have been adequately represented in the dataset.

## CONCLUSIONS

After a 5-year follow-up period, PVI as the first-line rhythm control therapy for AF demonstrated superior clinical outcomes compared to AAD, with lower risks of all-cause mortality, all-cause hospitalization, and heart failure exacerbation. These clinical benefits extended to paroxysmal and persistent AF subcohorts.

## Data Availability

Data are not available.

## SUPPLEMENTAL MATERIALS

**Supplemental Table 1.** Clinical Outcomes of Patients with Pulmonary Vein Isolation vs. Antiarrhythmic Drugs as First-Line Therapy for Paroxysmal Atrial Fibrillation (Before and After Propensity Score Matching).

|  | **PVI, n (%)** | **AAD, n (%)** | **Hazard Ratio**<br>**(95% CI)**<br>**(AAD as**<br>**reference)** | **P-value** |
| --- | --- | --- | --- | --- |
| **Primary Outcomes** |  |  |  |  |
| Composite Outcome | 218 (36.2) | 279 (46.3) | 0.71 (0.60 – 0.85) | <0.01 |
| All-Cause Mortality | 15 (2.5) | 31 (5.1) | 0.49 (0.27 – 0.91) | 0.02 |
| All-Cause Hospitalization | 192 (31.9) | 246 (40.9) | 0.71 (0.59 – 0.86) | <0.01 |
| Heart Failure Exacerbation | 69 (11.5) | 87 (14.5) | 0.79 (0.58 – 1.09) | 0.15 |
| **Secondary Outcomes** |  |  |  |  |
| Ischemic Stroke | 32 (5.3) | 30 (5.0) | 1.08 (0.66 – 1.78) | 0.76 |
| Major Bleeding Event (Intracranial or Gastrointestinal Bleeding) | 16 (2.7) | 31 (5.1) | 0.52 (0.28 – 0.95) | 0.03 |

**Supplemental Table 2.** Clinical Outcomes of Patients with Pulmonary Vein Isolation vs. Antiarrhythmic Drugs as First-Line Therapy for Persistent Atrial Fibrillation (Before and After Propensity Score Matching).

|  | **PVI, n (%)** | **AAD, n (%)** | **Hazard Ratio**<br>**(95% CI)**<br>**(AAD as**<br>**reference)** | **P-value** |
| --- | --- | --- | --- | --- |
| **Primary Outcomes** |  |  |  |  |
| Composite Outcome | 1,067 (42.9) | 1,291 (51.8) | 0.75 (0.69 – 0.81) | <0.01 |
| All-Cause Mortality | 106 (4.3) | 245 (9.8) | 0.42 (0.34 – 0.53) | <0.01 |
| All-Cause Hospitalization | 890 (35.7) | 1,063 (42.7) | 0.76 (0.69 – 0.83) | <0.01 |
| Heart Failure Exacerbation | 542 (21.8) | 634 (25.5) | 0.83 (0.74 – 0.93) | <0.01 |
| **Secondary Outcomes** |  |  |  |  |
| Ischemic Stroke | 158 (6.3) | 175 (7.0) | 0.89 (0.72 – 1.10) | 0.29 |
| Major Bleeding Event (Intracranial or Gastrointestinal Bleeding) | 112 (4.5) | 136 (5.5) | 0.81 (0.63 – 1.04) | 0.09 |

**Supplemental Figure 1.**
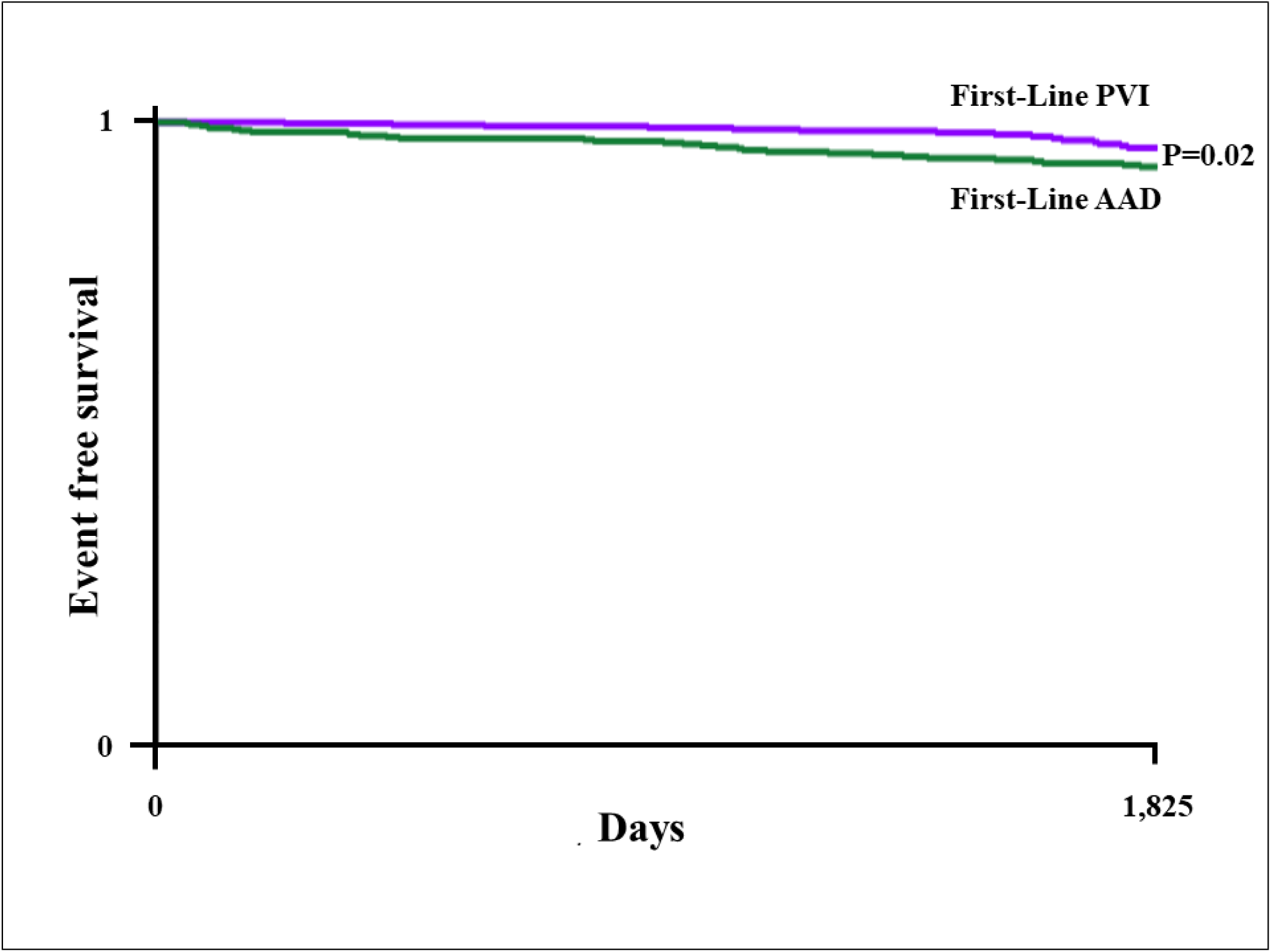
Kaplan-Meier Survival Analyses of All-Cause Mortality for Pulmonary Vein Isolation vs. Antiarrhythmic Drugs as First-Line Therapy for Paroxysmal Atrial Fibrillation.

**Supplemental Figure 2.**
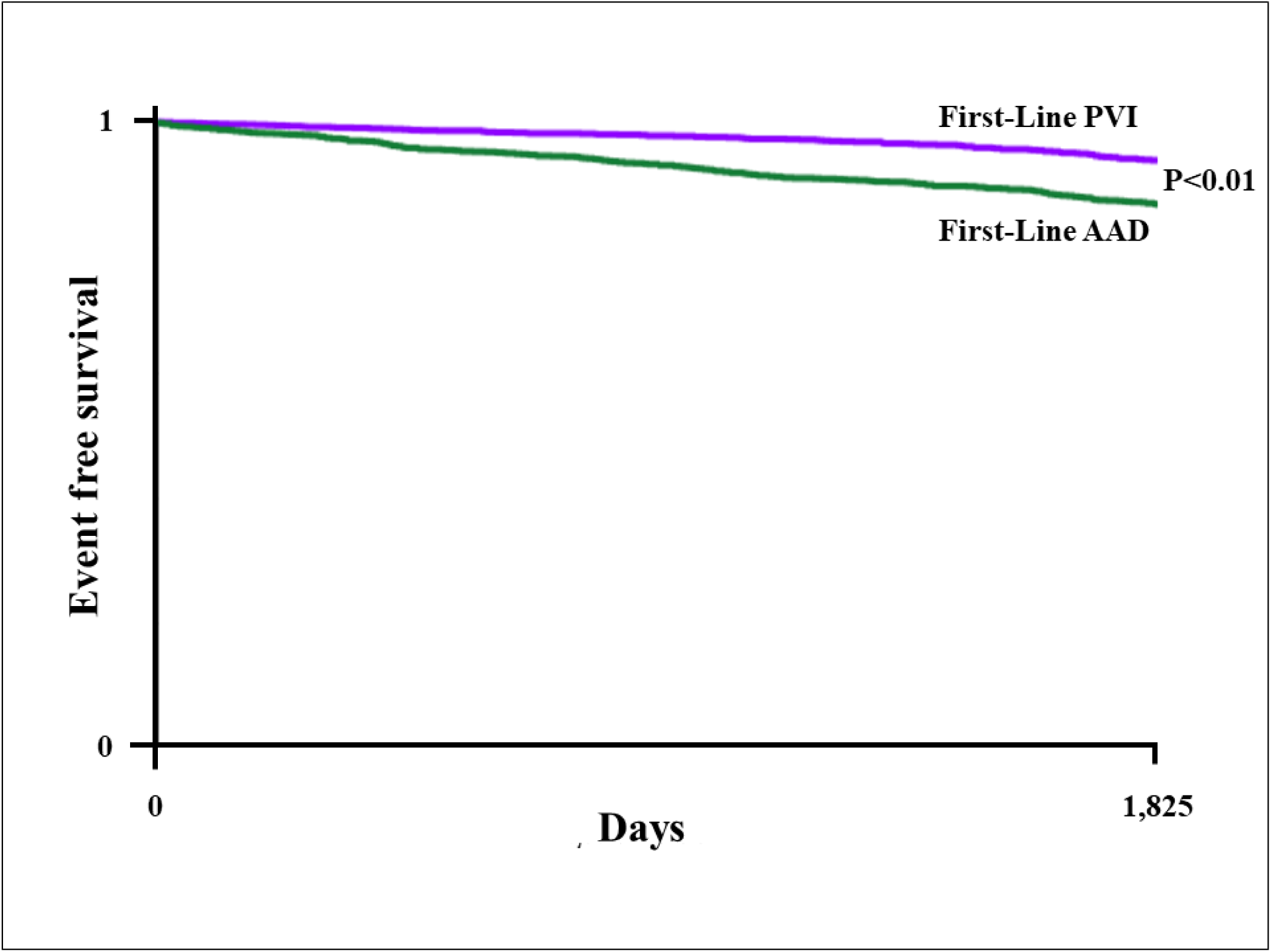
Kaplan-Meier Survival Analyses of All-Cause Mortality for Pulmonary Vein Isolation vs. Antiarrhythmic Drugs as First-Line Therapy for Persistent Atrial Fibrillation.

## REFERENCES

1. Elsheikh S, Hill A, Irving G, Lip GYH, Abdul-Rahim AH. Atrial fibrillation and stroke: State-of-the-art and future directions. Curr Probl Cardiol. 2024;49:102181. doi: 10.1016/j.cpcardiol.2023.102181

2. Lippi G, Sanchis-Gomar F, Cervellin G. Global epidemiology of atrial fibrillation: An increasing epidemic and public health challenge. Int J Stroke. 2021;16:217–221. doi: 10.1177/1747493019897870

3. Odutayo A, Wong CX, Hsiao AJ, Hopewell S, Altman DG, Emdin CA. Atrial fibrillation and risks of cardiovascular disease, renal disease, and death: systematic review and meta-analysis. BMJ. 2016;354:i4482. doi: 10.1136/bmj.i4482

4. Stabile G, Bertaglia E, Senatore G, De Simone A, Zoppo F, Donnici G, Turco P, Pascotto P, Fazzari M, Vitale DF. Catheter ablation treatment in patients with drug-refractory atrial fibrillation: a prospective, multi-centre, randomized, controlled study (Catheter Ablation For The Cure Of Atrial Fibrillation Study). Eur Heart J. 2006;27:216–221. doi: 10.1093/eurheartj/ehi583

5. Andrade JG, Wells GA, Deyell MW, Bennett M, Essebag V, Champagne J, Roux JF, Yung D, Skanes A, Khaykin Y, et al. Cryoablation or Drug Therapy for Initial Treatment of Atrial Fibrillation. N Engl J Med. 2021;384:305–315. doi: 10.1056/NEJMoa2029980

6. Cosedis Nielsen J, Johannessen A, Raatikainen P, Hindricks G, Walfridsson H, Kongstad O, Pehrson S, Englund A, Hartikainen J, Mortensen LS, et al. Radiofrequency ablation as initial therapy in paroxysmal atrial fibrillation. N Engl J Med. 2012;367:1587–1595. doi: 10.1056/NEJMoa1113566

7. Morillo CA, Verma A, Connolly SJ, Kuck KH, Nair GM, Champagne J, Sterns LD, Beresh H, Healey JS, Natale A, et al. Radiofrequency ablation vs antiarrhythmic drugs as first-line treatment of paroxysmal atrial fibrillation (RAAFT-2): a randomized trial. JAMA. 2014;311:692–700. doi: 10.1001/jama.2014.467

8. Wazni OM, Dandamudi G, Sood N, Hoyt R, Tyler J, Durrani S, Niebauer M, Makati K, Halperin B, Gauri A, et al. Cryoballoon Ablation as Initial Therapy for Atrial Fibrillation. N Engl J Med. 2021;384:316–324. doi: 10.1056/NEJMoa2029554

9. Writing Committee M, Joglar JA, Chung MK, Armbruster AL, Benjamin EJ, Chyou JY, Cronin EM, Deswal A, Eckhardt LL, Goldberger ZD, et al. 2023 ACC/AHA/ACCP/HRS Guideline for the Diagnosis and Management of Atrial Fibrillation: A Report of the American College of Cardiology/American Heart Association Joint Committee on Clinical Practice Guidelines. J Am Coll Cardiol. 2024;83:109–279. doi: 10.1016/j.jacc.2023.08.017

10. Kuniss M, Pavlovic N, Velagic V, Hermida JS, Healey S, Arena G, Badenco N, Meyer C, Chen J, Iacopino S, et al. Cryoballoon ablation vs. antiarrhythmic drugs: first-line therapy for patients with paroxysmal atrial fibrillation. Europace. 2021;23:1033–1041. doi: 10.1093/europace/euab029

11. Andrade JG, Deyell MW, Macle L, Wells GA, Bennett M, Essebag V, Champagne J, Roux JF, Yung D, Skanes A, et al. Progression of Atrial Fibrillation after Cryoablation or Drug Therapy. N Engl J Med. 2023;388:105–116. doi: 10.1056/NEJMoa2212540

12. Oraii A, McIntyre WF, Parkash R, Kowalik K, Razeghi G, Benz AP, Belley-Cote EP, Conen D, Connolly SJ, Tang ASL, et al. Atrial Fibrillation Ablation in Heart Failure With Reduced vs Preserved Ejection Fraction: A Systematic Review and Meta-Analysis. JAMA Cardiol. 2024;9:545–555. doi: 10.1001/jamacardio.2024.0675

13. Turagam MK, Musikantow D, Whang W, Koruth JS, Miller MA, Langan MN, Sofi A, Choudry S, Dukkipati SR, Reddy VY. Assessment of Catheter Ablation or Antiarrhythmic Drugs for First-line Therapy of Atrial Fibrillation: A Meta-analysis of Randomized Clinical Trials. JAMA Cardiol. 2021;6:697–705. doi: 10.1001/jamacardio.2021.0852

14. Marrouche NF, Brachmann J, Andresen D, Siebels J, Boersma L, Jordaens L, Merkely B, Pokushalov E, Sanders P, Proff J, et al. Catheter Ablation for Atrial Fibrillation with Heart Failure. N Engl J Med. 2018;378:417–427. doi: 10.1056/NEJMoa1707855

15. Packer DL, Mark DB, Robb RA, Monahan KH, Bahnson TD, Poole JE, Noseworthy PA, Rosenberg YD, Jeffries N, Mitchell LB, et al. Effect of Catheter Ablation vs Antiarrhythmic Drug Therapy on Mortality, Stroke, Bleeding, and Cardiac Arrest Among Patients With Atrial Fibrillation: The CABANA Randomized Clinical Trial. JAMA. 2019;321:1261–1274. doi: 10.1001/jama.2019.0693

16. Calkins H, Hindricks G, Cappato R, Kim YH, Saad EB, Aguinaga L, Akar JG, Badhwar V, Brugada J, Camm J, et al. 2017 HRS/EHRA/ECAS/APHRS/SOLAECE expert consensus statement on catheter and surgical ablation of atrial fibrillation. Heart Rhythm. 2017;14:e275–e444. doi: 10.1016/j.hrthm.2017.05.012

17. Joglar JA, Chung MK, Armbruster AL, Benjamin EJ, Chyou JY, Cronin EM, Deswal A, Eckhardt LL, Goldberger ZD, Gopinathannair R, et al. 2023 ACC/AHA/ACCP/HRS Guideline for the Diagnosis and Management of Atrial Fibrillation: A Report of the American College of Cardiology/American Heart Association Joint Committee on Clinical Practice Guidelines. Circulation. 2024;149:e1–e156. doi: 10.1161/CIR.0000000000001193

18. Sjalander S, Holmqvist F, Smith JG, Platonov PG, Kesek M, Svensson PJ, Blomstrom-Lundqvist C, Tabrizi F, Tapanainen J, Poci D, et al. Assessment of Use vs Discontinuation of Oral Anticoagulation After Pulmonary Vein Isolation in Patients With Atrial Fibrillation. JAMA Cardiol. 2017;2:146–152. doi: 10.1001/jamacardio.2016.4179

